# Living with Long COVID: Implementing a living approach to the NICE guideline on managing the long-term effects of COVID-19

**DOI:** 10.1101/2023.05.09.23289572

**Authors:** Steve Sharp, Sarah Boyce, Justine Karpusheff, Fiona Glen

**Author notes:** **Address for correspondence:**, NICE, Level 1A, City Tower, Manchester, M1 4BT, United Kingdom. **Authors’ contributions** Steve Sharp: Conceptualisation, Writing - Original Draft, Writing - Review & Editing. Sarah Boyce: Writing - Original Draft, Writing - Review & Editing. Justine Karpusheff: Writing - Original Draft, Writing - Review & Editing. Fiona Glen: Writing - Review & Editing.

## Abstract

**Objectives:** The aim of this paper is to describe the development, implementation and evaluation of a flexible living approach to maintaining NICE’s long-term effects of COVID-19 (LTE) guideline and monitoring the uncertain evidence base of this condition.

**Study Design and Setting:** The NICE COVID-19 team reviewed its practical experiences of establishing a living approach to developing and maintaining the LTE guideline, including initial development, maintenance and eventual transition to a lower intensity model. The methods and processes were described narratively over the first 2 years of the guideline’s lifespan. This was combined with quantitative data on emerging and cumulative evidence over the period to chart the evidence landscape.

**Results:** Following publication, the initial timepoint-based update process evolved into a flexible living approach with remote topic expert engagement.

Experts engaged with the new process with a 64% response rate to the online surveys.

Emerging evidence increased rapidly following publication [11,405 studies assessed in 2021 and 13,181 in 2022] and was captured by continuous surveillance. There were no urgent triggers for updating from the studies identified in 2022 via the living approach, saving considerable resources over the timepoint based approach which would commit resources to planning and convening expert panel meetings.

A total of 184 studies with a potential future impact were summarised to capture the cumulative evidence base. Experts highlighted ongoing research and implementation issues which have further informed surveillance of the guideline.

After a sustained period without triggers for updating, the living approach was restricted to the highest priority areas with surveillance of ongoing studies.

**Conclusion:** This paper illustrates a flexible living approach taken to a novel condition with an evolving evidence landscape. Currency of some living guidelines can be maintained without the need for frequent updating.

**Highlights: What is new?:**

- In an unpredictable pandemic context, novel conditions with uncertain aetiology, diagnosis, management and prognosis demand a flexible living approach to surveillance of initial recommendations, even where triggers for updating remain infrequent.
- Monitoring cumulative evidence with potential future impact is important for high priority areas lacking a strong evidence base.
- In guidelines with previous scheduled updates, transition to a more reactive ‘trigger-based’ approach can be both more efficient and productive, while maintaining currency of recommendations through continuous surveillance.
- Determining when to transition between living and standard approaches to maintaining a guideline is dependent on multiple factors, including intelligence from the health and social care system, ongoing research and government policy.

**Use of NICE COVID-19 content internationally:** Our COVID-19 rapid guidelines and evidence summaries are exempt from our overseas reuse application, licence and fee. This means you can:

- adopt the guidelines for your own healthcare setting
- adapt the guidelines by combining them with your own local content
- translate the resultant outputs.

When using content from our COVID-19 rapid guidelines and evidence summaries you must:

- make all your outputs reusing NICE content freely available to others
- acknowledge the use of NICE content, and link to the source content on our website
- only use the NICE logo if the original NICE guidance publication is used in its entirety without including additional content
- tell us how our content has been used by emailing, to support the evaluation and development of our guidance.

We cannot accept responsibility or liability for the use of our content in third party outputs.

Further information on reuse of content is available on the NICE website.

## 1. Introduction

Since March 2020, NICE has been producing rapid guidelines on COVID-19 to support the healthcare system during the pandemic. Six months into the pandemic, NICE’s 24th rapid guideline, Managing the long-term effects (LTE) of COVID-19 (NICE, 2020) was developed within a compressed timeframe in response to the increasing prevalence of this emerging condition of diverse, persisting symptoms following the acute phase of the disease. An immediate challenge lay in how to approach the surveillance of the published guideline with a growing but largely uncertain evidence base, with many knowledge gaps and evidence emerging at different rates across sup-topics. This variability has presented challenges in optimising the maintenance of the guideline and has signalled the need for a flexible model that is responsive to emerging evidence in key areas.

Despite the World Health Organization (WHO) announcing in May 2023 that COVID-19 is no longer a public health emergency of international concern (WHO, 2023), the most recent estimates indicate that 1.9 million people in private households in the UK (2.9% of the population) were still experiencing self-reported long COVID (ONS, 2023). This underlines the ongoing need to provide timely and relevant guidance to the health and care system.

Aim: to describe the development and implementation of a flexible living approach to maintaining the currency of the LTE guideline and managing the uncertain evidence base of this condition.

This information is useful in informing planning and operations for future living guideline topic areas.

## 2. Methods

We undertook a retrospective analysis of the guideline development, surveillance and updating stages from its initial development and publication in December 2020 through to a reactive ‘trigger-based’ living approach adopted in 2022, documenting:

- Original and revised methods and processes
- Point-in-time and trends over time data on emerging and cumulative evidence
- Web page visits during the lifespan of the guideline
- Remote and face to face expert engagement

Methods and processes used in the development of the guideline and any revisions during surveillance and updating were reviewed. The rationales for the evolving methods were explored in the context of the expanding evidence base and other contextual factors, including resource use, topicality, level of uncertainty and urgency of system need.

Data on the volume of evidence was obtained from the NICE COVID-19 evidence repository; a database of cumulative studies stored in EPPI Reviewer 5 software that includes all search results from when surveillance searches for the COVID-19 pandemic began in March 2020. The repository section for LTE was used to capture the new and cumulative evidence covering the 2020 initial development; the 2021 evidence landscaping, re-scoping and update; and for the 2022 flexible living approach.

Historical web page visits to the guideline pages were obtained from the NICE Digital, Information and Technology function as an indicator of guideline usage.

Expert engagement data was obtained on frequency and duration of panel meetings convened during initial development and first update, and from online survey data of the flexible living approach.

## 3. Results

### 3.1. 2020 Topic referral and guideline development

The key developments from topic referral in October 2020 to publication are summarised in Box 1.

#### Box 1.

Key developments: Version 1 2020

- Following topic referral a 12 week development period was adopted
- Collaborative approach to guideline development established with SIGN and RCGP
- Scope of guideline agreed, including case definition and coding
- Acknowledgement of a sparse evidence base led to the use of real world evidence and expert testimony
- Modified methods of development included targeted stakeholder consultation
- High levels of interest, with extensive media coverage and debate
- Established agreements to share evidence with international guideline producers working on the same topic
- Developed research recommendations that fed into NIHR funded call for research

#### 3.1.1 Referral and initial development

The prolonged effects of acute COVID-19 manifesting as a distinct condition first appeared in summer 2020, several weeks into the pandemic when a sub-group of people first experiencing the acute disease reported persisting symptoms several weeks after onset (Tenforde, 2020). An increasing and unknown epidemiology of these sequelae led to an urgent NICE referral from NHS England to produce the guideline.

Previous NICE rapid guidelines had been developed in single week timeframes with more limited scopes and minimal evidence. It was recognised that to better balance rigour, coverage and urgency there was a need to extend the development timeline. This ensured a full review of the emerging evidence base and enabled stakeholder involvement, an area highlighted by a rapid review as missing from the development process in some emergency guidelines, because of the need for expedited guidance (Dagens, 2020).

The guideline was developed with external partners Scottish Intercollegiate Guidelines Network (SIGN) and Royal College of General practitioners (RCGP) to ensure the guideline adopted a UK wide perspective and acknowledged the vital role primary care services needed to play in caring for people with this condition. This collaboration included operational and strategic decision-making throughout development, with evidence reviews allocated between organisations and frequent oversight meetings.

#### 3.1.2 Scoping and evidence identification

An expert advisory panel was formed and convened to discuss and agree the guideline scope. Key sections were agreed to reflect the clinical pathway and service organisation. There was a strong consensus among experts and stakeholders to develop a clinical case definition and clinical diagnostic and referral codes, without which diagnosis would be problematic and difficult to record. This was a departure from usual guideline production, but it was acknowledged that this would support implementation of the recommendations and enable data on an emerging condition to be gathered to further inform recommendations. Once definitions and codes were produced, the NICE team produced review questions and protocols for each area followed by a systematic search for published evidence (Levay and Finnigan, 2021).

In addition to questions on how to treat the condition, the uncertainty in the evidence base also extended to its pathological mechanisms as well as patient characteristics and how their experiences might inform the development of new service models. As such, a convergent mixed methods approach (Stern, 2020) was adopted in developing review questions and reviewing the evidence which enabled the panel to explore how far people’s lived experiences supported or challenged the limited quantitative data. This approach also improved the understanding of how healthcare might support people experiencing LTE, and to determine the nature of the condition and how it is experienced.

Since the condition had only recently emerged without a clearly defined case definition, the evidence base was in its infancy and was characterised by consensus based guidance, service delivery case studies and small, often uncontrolled, observational studies without clearly defined populations. With low quality evidence, the gaps in research on service delivery were addressed by extending evidence sources to expert witnesses who could provide testimony based on clinical experience from real world clinical settings. Real world evidence was also used from the Zoe <u>COVID-19 symptom tracker mobile application</u> a free mobile phone app developed by ZOE Limited and King’s College London (Menni et al., 2020) which enabled users to self-report symptoms.

#### 3.3.3 Validation

To accelerate publication, the guideline was subject to a compressed, targeted stakeholder consultation lasting 1 week, a shorter duration than the normal 4-6 week period. A range of stakeholders were invited to take part, including relevant national professional, and patient or carer groups. Consultation comments were collated in a thematic summary with responses agreed with the panel.

Following the pragmatic quality assurance checks and review done iteratively throughout guideline development on both the guideline and evidence reviews, the guideline was approved by senior members of the development teams at NICE, SIGN and the RCGP.

#### 3.3.4 Post-publication dissemination

Publication led to a high level of media and public interest, with over 100,000 website hits in the 2 weeks following publication and widespread press coverage. NICE disseminated the guideline nationally and internationally via a collaborative group of guideline producers covering several countries. In some cases, reciprocal arrangements were made in the evidence ecosystem (Ravaud, 2020) by NICE sharing surveillance post-publication evidence with organisations performing living systematic reviews. By providing the most up to date surveillance results, economies were achieved in maintaining the currency of living reviews, such as through collaboration with the Public Health Agency of Canada (Domingo et al., 2021.

Further work with the National Insitute for Health Research (NIHR) informed the calls for research on the topic, by making research recommendations to prioritise evidence gaps. This led to numerous NIHR funded research projects, including <u>PHOSP-COVID</u> (PHOSP Consortium, 2023) <u>HEAL-COVID</u> (Liverpool Clinical Trials Consortium, 2023) and <u>STIMULATE-ICP</u> (University College London, 2023). The research recommendations were driven by the high level of importance to patients, the national priority of Long COVID and its potential impact on the health and care system.

### 3.2. 2021 Surveillance and update

#### Box 2.

Key developments: 2021 Initial surveillance/evidence landscaping

- Weekly surveillance was established to monitor new evidence
- Proliferation of new studies but low quality and no strong signals for updating
- Changing evidence landscape exposed gaps in evidence for vaccine impact; children and young people; and barriers and facilitators to referral
- NICE committed to a timepoint based update in 2021 based on gap analysis and re-scoping
- Inconsistent coding and research classification highlighted the need to incorporate case definition into recommendations

#### 3.2.1. Evidence landscaping and re-scoping

Following publication, NICE committed to a living approach comprising continuous weekly surveillance of the rapidly expanding evidence base (Figures 1 and 2 show new emerging evidence and the cumulative evidence base, respectively). However, this exposed a lack of coordinated research as a barrier to an efficient evidence ecosystem, with many low quality studies often duplicating the conclusions of existing evidence. This was particularly apparent in studies of prevalence, signs and symptoms, by far the most prolific area of research which was rapidly approaching saturation point without adding value to the knowledge base. This eventually led to refinement in inclusion criteria by restricting this area to living systematic reviews and the largest primary studies.

**Figure 1.**
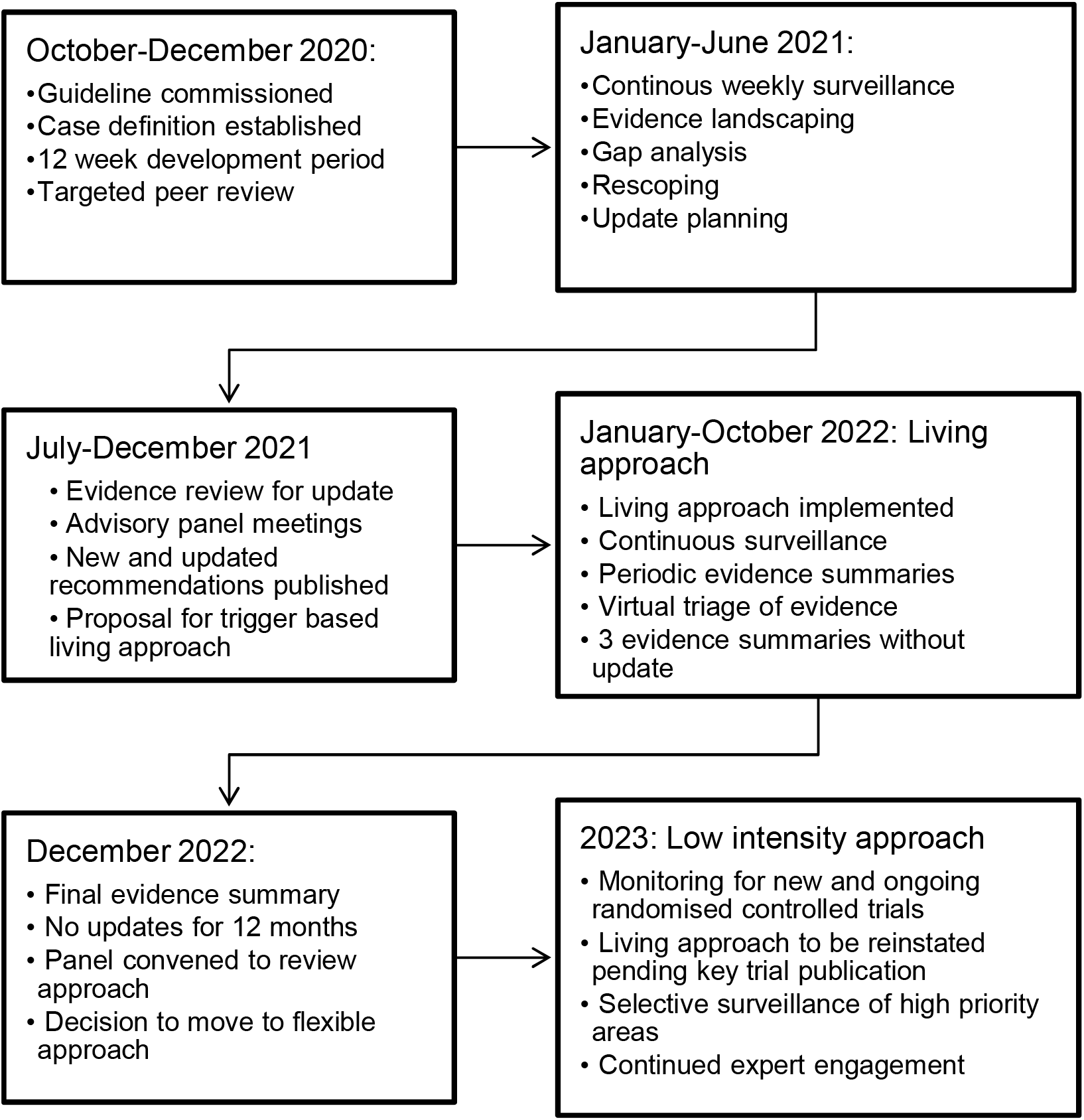
Timeline of guideline.

**Figure 1.**
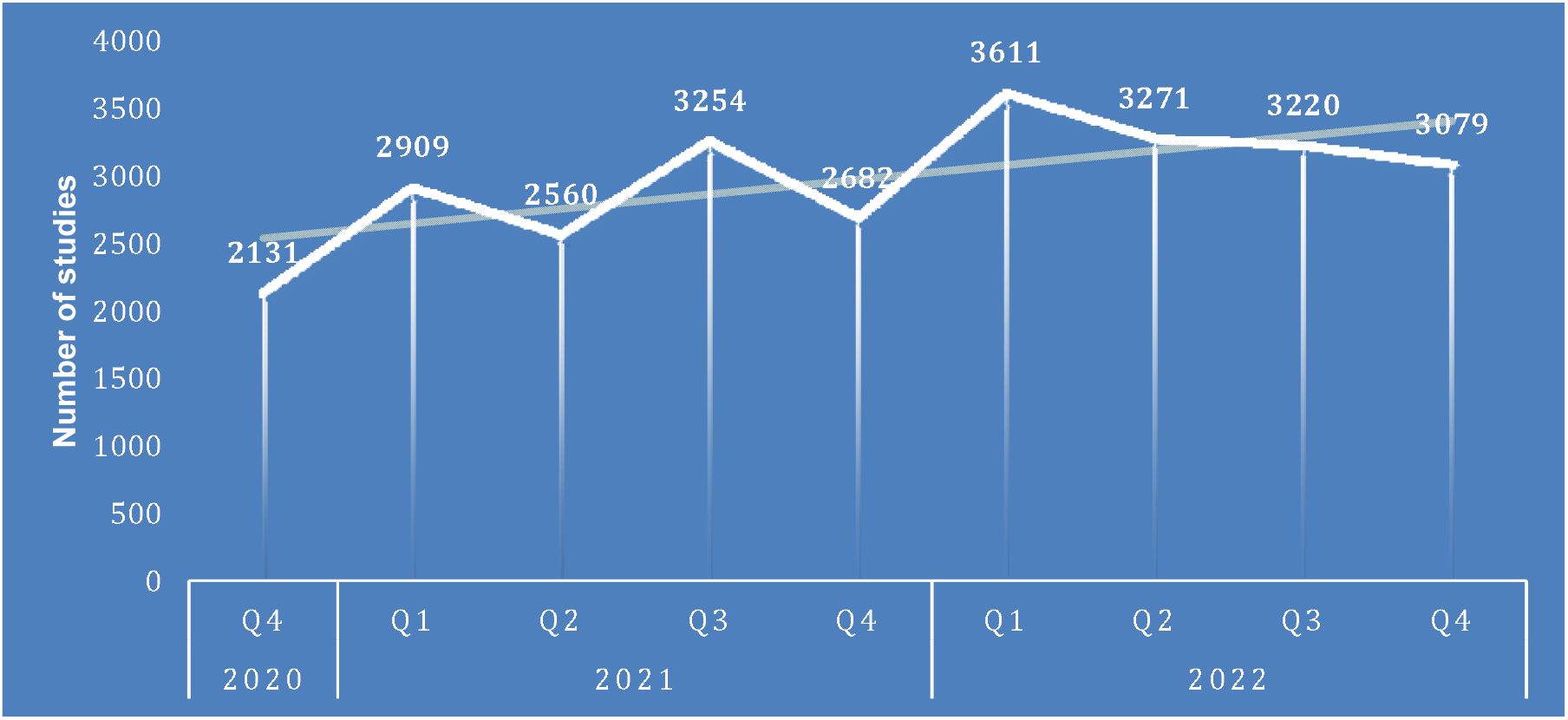
New studies identified by quarter 2020-2022.

**Figure 2.**
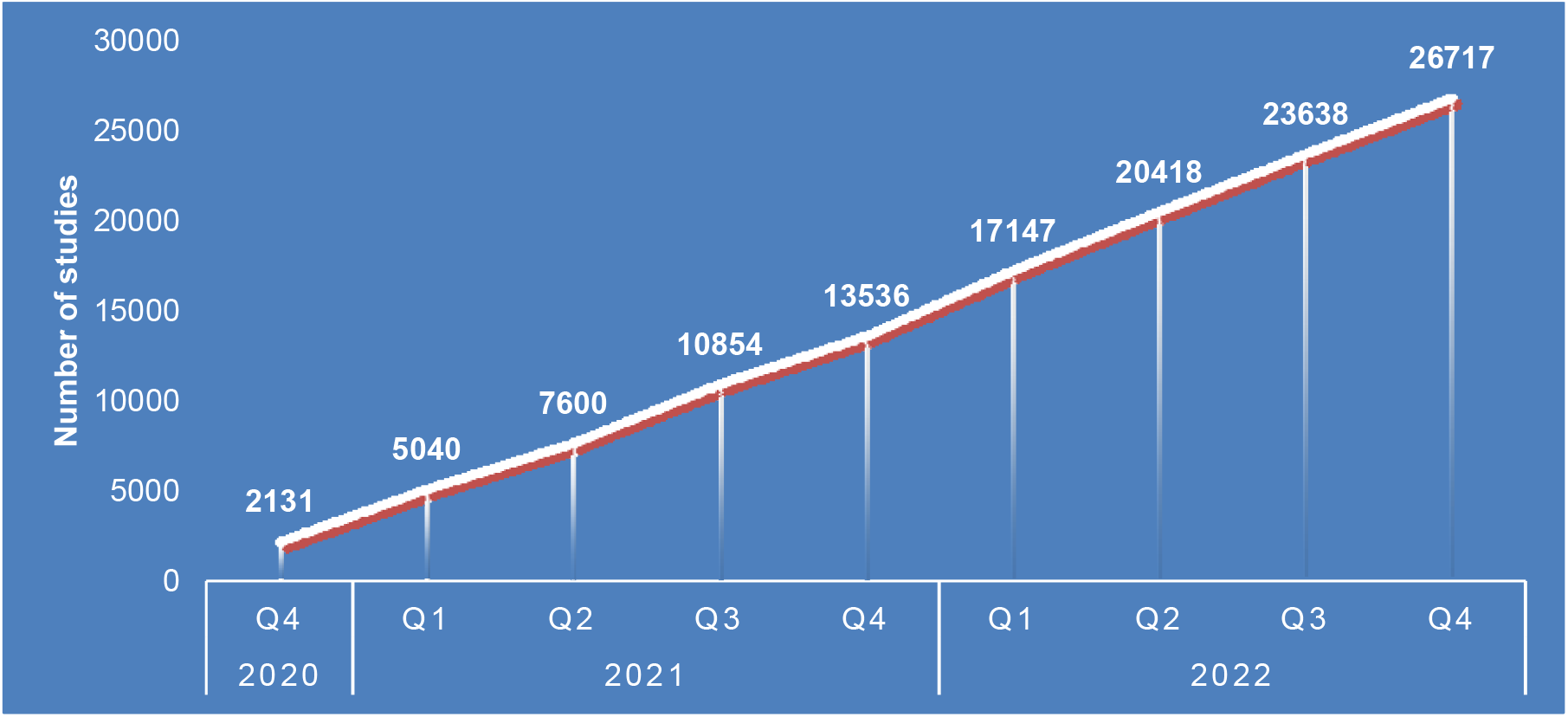
Cumulative studies identified by quarter 2020-2022.

The early surveillance identified gaps in the guideline, particularly with the introduction of vaccination and the associated uncertain impact on LTE. Evidence on age sub-groups particularly children and young people was also lacking. Early analysis of electronic health records (Walker et al., 2021) also revealed the variable and inconsistent clinical coding across England, suggesting low levels of awareness of the case definition. These findings led to a re-scoping exercise to incorporate these key areas into the scope with stakeholder input and agreement, followed by a scheduled update to the guideline. The stakeholder workshop ensured that revisions to the scope were driven by evidence but were still informed by the diverse stakeholder perspectives. This was managed through objective presentation of the evidence landscape, focus groups via breakout rooms and group discussion and agreement.

#### 3.2.2. First update

##### Box 3

Key Developments: 2021 update

- Re-scoping with stakeholder input included case definition review question, impact of vaccines, children and young people, referral barriers
- Guideline content was migrated to an online authoring platform to enable a more responsive and transparent approach to updating
- Update proceeded and was published in November 2021 based on evidence to June 2021
- Limited updating from the resource intensive approach led to a process review
- The need for a more flexible trigger-based approach was identified

The re-scoping process extended the timescale of the scheduled update to the guideline, but this was still completed rapidly after a series of panel meetings had been compressed into a short intense period. This underlined the importance of gaining expert commitment to providing frequent input from the outset of a living guideline. Indeed, expert engagement presented challenges throughout the lifecycle of the guideline with all meetings convened remotely and in rapid succession before guideline publication. The short development timeframe imposed trade-offs around accelerating the meeting preparation and follow-up tasks, such as the continuous maintenance of the declarations of interests register, which was often updated at short notice ahead of meetings. However, transparency of decision-making and reporting was maintained in line with NICE’s standards, which was also facilitated by the use of the web-based guideline authoring platform Making GRADE the Irresistible Choice (MAGICapp, 2023) https://magicevidence.org. This has the facility to allow annotation of recommendations with the use of tags to show, for example, new and updated recommendations. It also allows the addition of rationale text to individual recommendations. Use of transparent presentation of changes made in a living approach allows the reader to understand where changes have been made and why.

One of the key learning points of the early living approach was that scheduled point-in-time updates to a guideline can be resource intensive where the update extends to an expanded scope of a topic area with significant knowledge gaps. Review of the evidence revealed only limited new evidence in a few areas which did not significantly challenge the current recommendations. Although the scope revisions were necessary, to incorporate new areas emerging since the original publication, some areas were clearly more dynamic in generating new evidence than others. This indicated that updating at a specific timepoint was not the most efficient or productive approach, despite proliferating evidence, and signalled the need for a different approach to future surveillance and updating.

### 3.3. 2022 Trigger-based living approach

#### Box 4

Key developments: 2022 Flexible living approach

- A continuous trigger-based approach was launched to respond to emerging evidence
- Urgent sections of the guideline were prioritised for potential updating via remote triage without the need to commit resources to definite updates
- High levels of emerging evidence, political interest and ongoing uncertainty underlined the need for continuous monitoring
- Studies with potential future impact were captured to monitor cumulative evidence over time
- Evidence summaries were circulated to the expert panel for assessment of evidence and remote submission of living intelligence
- New and cumulative evidence was skewed towards lower priority areas, with research gaps ongoing
- A lack of update triggers led to the transition to a low intensity model with a mechanism for future fast track updating

#### 3.3.1 Continuous living approach

Following the update a more flexible living approach was adopted, comprising continuous monitoring of emergent evidence and a quarterly summary of the evidence landscape. This was circulated to the expert advisory panel with an online survey to ensure that both new and cumulative evidence was considered in the need to update, alongside the experts’ input and intelligence. This remote triage process avoided the need to commit resources to convening panel meetings unless these were deemed necessary by evidence with impact.

#### 3.3.2 Remote expert engagement

During 2022 some potential triggers for updating emerged, such as in the impact of vaccines and the use of screening tools, which divided expert opinion to some extent but with the majority still considering the guideline to be current. This underlines the value of topic expert input in assessing thresholds for updating, particularly where evidence is more nuanced than in traditional intervention trials. To this end, the quarterly online surveys were instrumental in the living approach by facilitating expert ‘triage’ of evidence as well as providing health system intelligence on local implementation of the guideline, equalities issues and ongoing research.

Over the course of the maintenance phase of the living guideline, it became clear that many recommendations in the guideline needed to remain broad and all-encompassing rather than prescriptive in nature, because the condition is individually specific with disparate symptoms and trajectories.

#### 3.3.3 Transition from high intensity approach

After 12 months of the trigger based approach without strong triggers for updating, the guideline entered the transition phase from the high intensity living mode to a lower intensity approach of horizon scanning for ongoing research and intelligence gathering through periodic surveys of the expert advisory panel. To ensure that the guideline will remain responsive to emergent evidence, a mechanism has been set up to enable accelerated updating as key studies publish.

## 4. Discussion

A distinguishing feature of living guidelines is frequent updating of individual recommendations or sections in the light of new evidence. However, other drivers may justify a living approach, including urgency of system need, high political and public interest (over 1 million web page visits between 2020-2022, averaging approximately 15,000 visits per month during 2022)[NICE: web statistics 2020-2022], and the ongoing uncertainty of managing a novel condition. This applies to LTE where continuous surveillance to demonstrate guideline currency in the light of cumulative evidence has been necessary even without frequent updating. We have shown that the rapidly expanding evidence base (Figures 1 and 2) presented a resource intensive challenge in maintaining currency but is feasible using innovative methods of surveillance and expert engagement.

Unpredictability during a pandemic demands vigilance in a living guideline context, where circumstances can change rapidly. Expedited trials of new and repurposed drugs, the emergence of vaccines and new variants all drive the need to embed practice changes quickly. Conversely, evidence can take longer periods to emerge for conditions such as LTE where uncertain pathological mechanisms and treatable traits lead to research hesitancy. Flexibility in living guideline methods therefore becomes crucial in efficient use of resources and we have shown that a guideline can be maintained with various models depending on the stage of the life cycle and the prevailing evidence landscape.

Our case study has several strengths. Firstly, it captures the evolution of a living guideline on a novel condition with high levels of uncertainty, urgent system need and rapid rate of change. It also presents a model for flexible surveillance and updating of a guideline that optimises use of resources. The constant review and refinement of methods has shown the value of an adaptable approach to monitoring the evidence base which may be transferable to other topics. We have also charted the journey of a topical guideline through all stages of the living guideline cycle comprising initiation, maintenance and retirement from living mode. The limitations of our case study include the focus on a single COVID-19 specific topic area, and we recognise that the high volume of research generated by the pandemic may not be generalisable to all non-COVID topics. However, the study contributes significantly to the body of evidence on approaches to living guidelines. Further research is needed on the value of this approach across a broader sample of topics.

## 5. Conclusion

Adopting a living approach to conditions with a high level of uncertainty, dynamic evidence base and urgent system need does not always necessitate frequent updates. Adapting the living approach within the changing evidence landscape allows efficient use of resources and timely assessment of new evidence.

## Data Availability

All data produced in the present study are available upon reasonable request to the authors

## Acknowledgements

The authors would like to thank the SIGN, RCGP and NICE COVID-19 teams for collaborating on the development and maintenance of the guideline throughout its lifecycle.

